# Analysis of Epidemiological Characteristics of Varicella in a Suburban Area of Pudong New District, Shanghai from 2011 to 2022: A Retrospective Study

**DOI:** 10.1101/2025.09.10.25335482

**Authors:** Li Yong, Liu Bo, Huang Xi-ya

**Affiliations:** Department of Public Health, Gaoqiao Community Health Service Center, Pudong New District, Shanghai,China; Institute of Infectious Disease Control and Prevention, Shanghai Municipal Center for Disease Control and Prevention, Shanghai,China; Pudong New District Center for Disease Control and Prevention (Pudong New District Health Supervision Institute), Shanghai,China

**Keywords:** Varicella, Epidemiological analysis, Prevention, control measures

## Abstract

**Objective:** To understand the incidence of varicella in a suburban area of Pudong New District, Shanghai, and provide a scientific basis for relevant prevention and control work.

**Methods:** Descriptive epidemiological methods were used to describe and analyze all reported varicella cases with the residential address in a suburban area from 2011 to 2022. SPSS software was used for statistical processing, and the chi -square test was used for the comparison of rates.

**Results:** A total of 1,708 varicella cases were reported in a suburban area from 2011 to 2022, with an average incidence rate of 107.72 per 100,000. The incidence showed obvious seasonal changes, presenting a “bimodal distribution”, with peaks from May to July and in November. The onset age was concentrated in the 1 - 14 age group, accounting for 54.27% of all cases. The affected population was mainly students (accounting for 37.30%), kindergarten children (accounting for 15.11%) and scattered children (accounting for 13.00%), and there were more males than females.

**Conclusion:** Students and kindergarten children are the key populations for varicella prevention and control. Epidemic monitoring should be strengthened during the peak incidence season every year, and the work of varicella vaccination and booster vaccination should be improved to comprehensively reduce the incidence of varicella.

---

Varicella is an acute viral infectious disease caused by varicella - zoster virus, which is transmitted through the respiratory tract or direct contact and is characterized by systemic herpes. It is generally transmitted through airborne droplets, contact with fresh vesicle fluid of patients or mucosal secretions, and is a common infectious disease in children. The main symptoms are fever accompanied by itchy rashes, papules and blisters all over the body ^[1 - 2]^. It is more common in children, highly contagious, and usually shows symptoms 10 - 21 days after infection ^[3]^. It is easy to cause small - scale epidemics, and lifelong immunity can be obtained after recovery ^[4]^. Varicella has obvious seasonality, being more common in winter and spring. It is highly infectious, spreads quickly, and is prone to cause outbreaks. It is a common infectious disease among children in schools, kindergartens and other institutions, seriously affecting children’s health, study and life, and has become a major public health problem ^[5 - 6]^. To understand the epidemiological characteristics and trends of varicella in a suburban area of Pudong New District, Shanghai, and provide a basis for formulating scientific varicella prevention and control measures, the varicella epidemic in a suburban area from 2011 to 2022 was analyzed.

## Materials and Methods

### Data Sources

The data on the varicella epidemic is sourced from the “China Disease Prevention and Control Information System”. Starting from May 10, 2025, the data is statistically analyzed based on conditions such as “date of onset” and “current residence address”. The statistical period for the date of onset is from January 1, 2011 to December 31, 2022. The population data is from the Gaohao Town Police Station in Pudong District, Shanghai.

### Statistical Methods

The epidemic data were exported from the “China Disease Prevention and Control Information System” according to the set conditions. WPS2016 was used to collect data, and descriptive epidemiological analysis was carried out on the three - dimensional distribution of varicella.

### Statistical Analysis

The data were analyzed and processed using WPS2016 and SPSS 23.0. The chi - square test was used for the comparison of rates, with the test level of α = 0.05. A P - value less than 0.05 was considered statistically significant.

## Results

### Incidence Overview

From January 1, 2011, to December 31, 2022, a total of 1,708 varicella cases were reported in a suburban area. The annual incidence fluctuated between 35.46 per 100,000 and 227.82 per 100,000, and the average annual incidence was 107.72 per 100,000. The year with the highest incidence was 2017, and the lowest was 2022. There was a statistically significant difference in the incidence among years (χ^2^ = 356.308, P < 0.05). There were no death cases among all the cases. The reported incidence showed a wave - like pattern of alternating high and low every year, with an obvious peak in 2017, and then gradually began to decline. See Figure 1 for the case reporting situation.

**Fig. 1.**
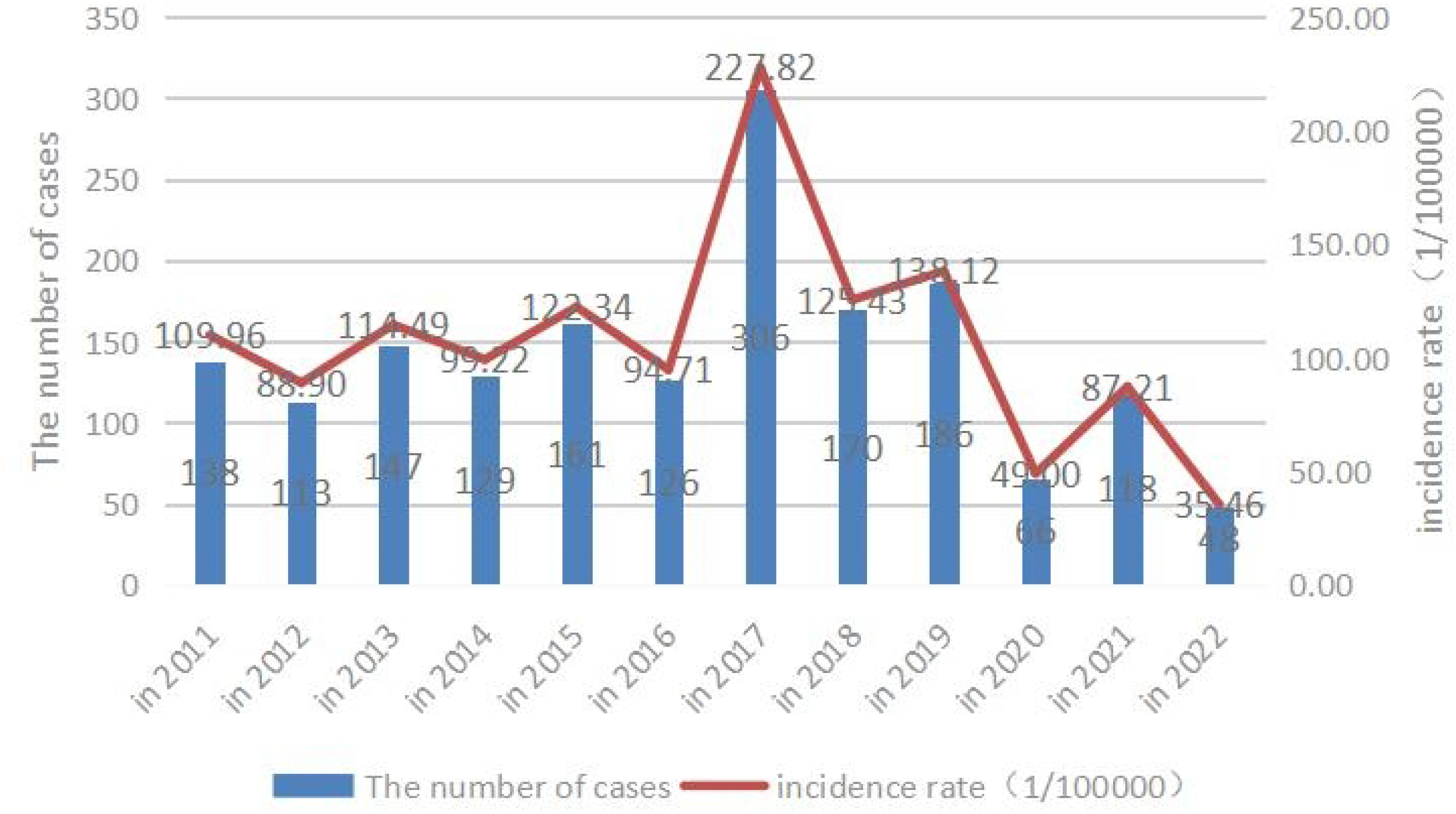

### Epidemiological Characteristics

#### Gender Distribution

Among the 1,708 reported varicella cases in a suburban area from 2011 to 2022, there were 917 male patients, accounting for 53.69%; and 791 female patients, accounting for 46.31%. The male - to - female ratio was 1.16:1. The incidence rate of males was 115.27 per 100,000, and that of females was 99.99 per 100,000. The incidence rate ratio was 1.15:1, and there was a statistically significant difference in gender (χ^2^ = 29.89, P < 0.05). See Figure 2 for the gender - specific incidence rates.

**Fig. 2.**
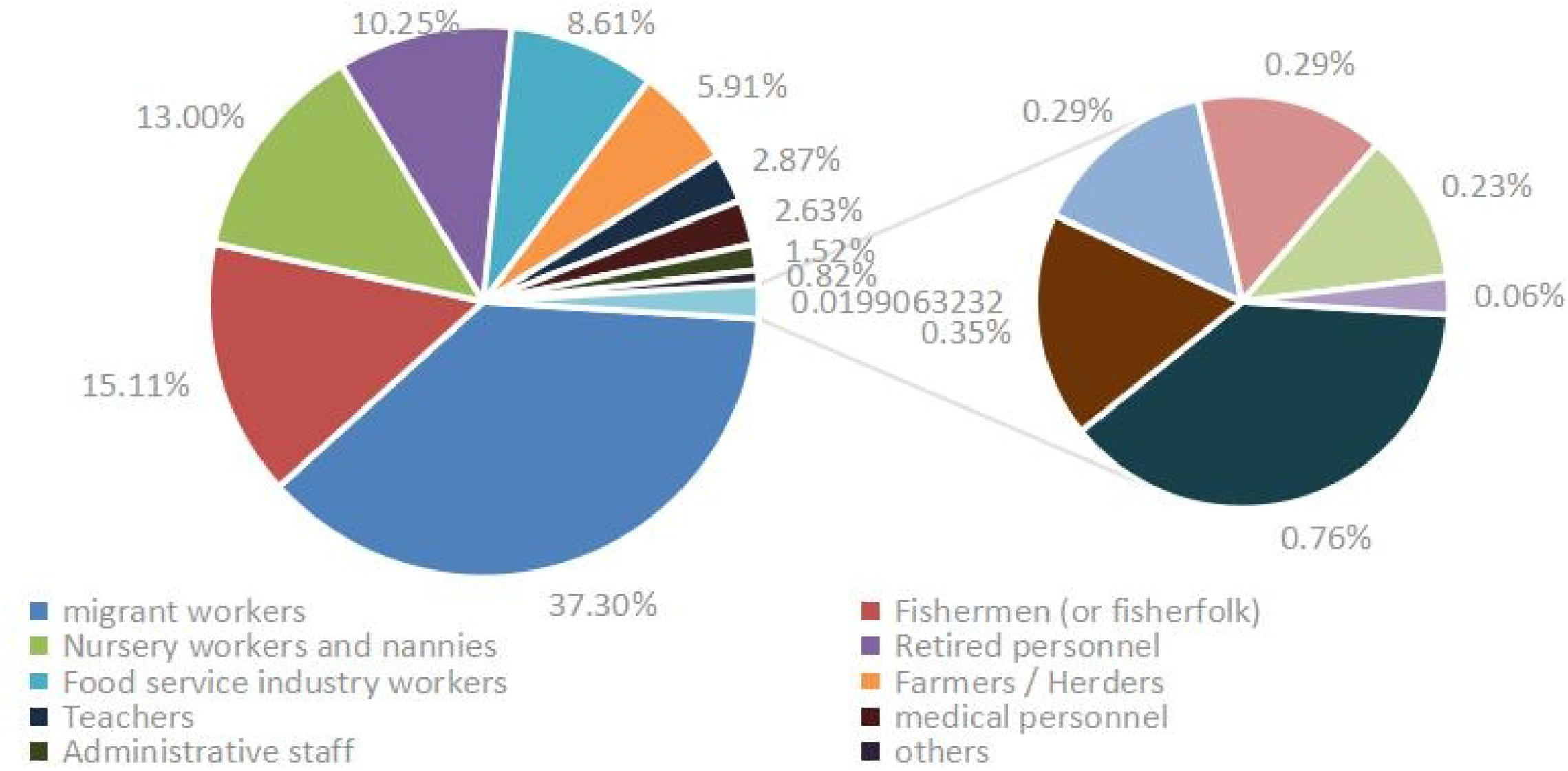

#### Age Distribution

According to the actual situation of the suburban area, age grouping was carried out. It was found that from 2011 to 2022, the age group with the most varicella cases was 5 - 9 years old, with a total of 406 cases, accounting for 23.77% of the total number of cases; followed by the 10 - 14 age group, with a total of 266 cases, accounting for 15.57% of the total number of cases; and the third was the 1 - 4 age group, with a total of 255 cases, accounting for 14.93% of the total number of cases. The youngest onset age was 28 days, and the oldest was 99 years old. The top three age groups in terms of incidence rate were: 5 - 9 years old (585.81 per 100,000), 1 - 4 years old (478.06 per 100,000), and <1 year old (438.37 per 100,000). There was a statistically significant difference in age (χ^2^ = 4719.431, P < 0.05). See Figure 3 for the incidence rates of each age group (per 100,000).

**Fig. 3.**
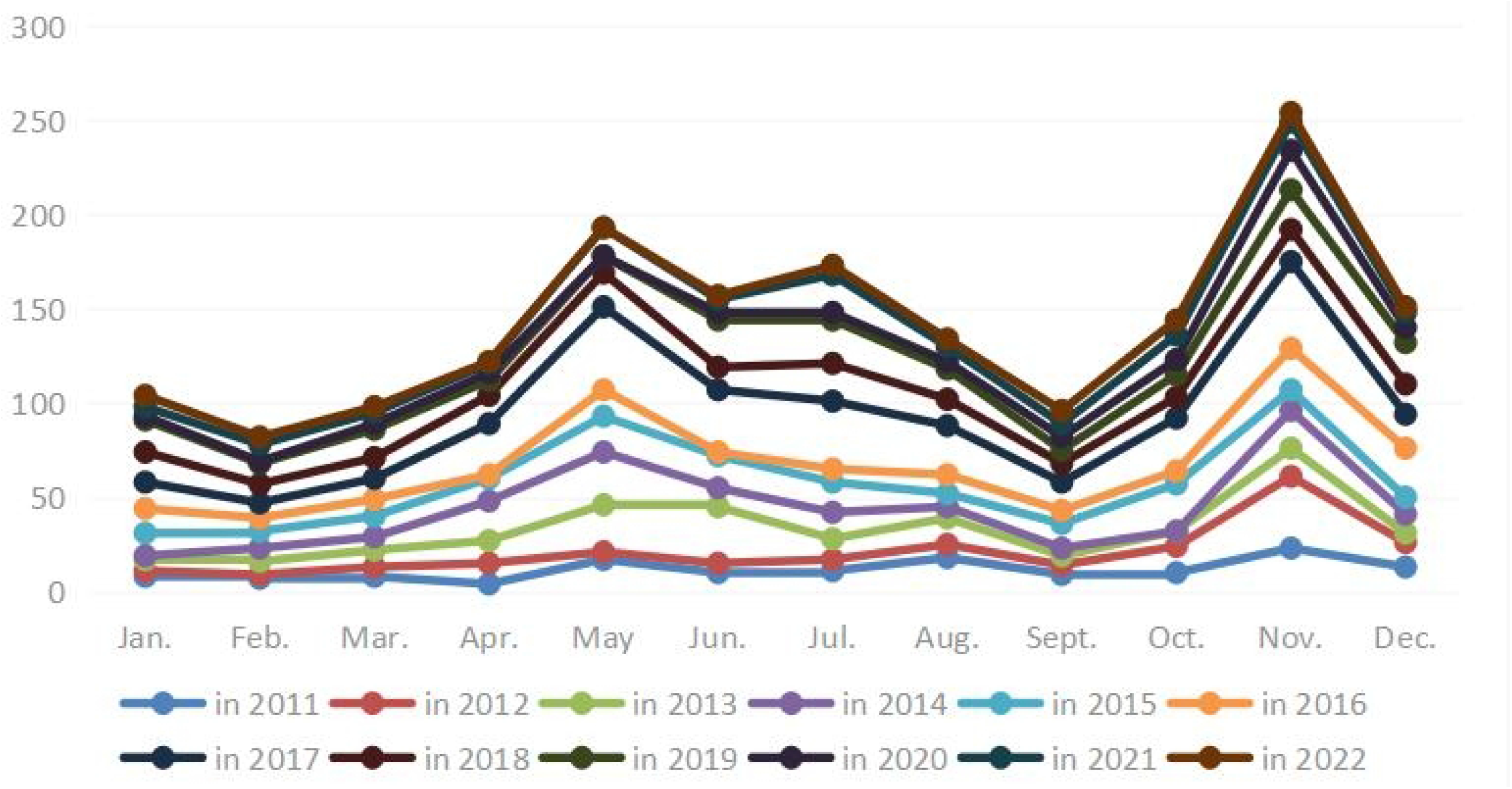

#### Occupation Distribution

The patients were mainly distributed in 17 occupations, including students, kindergarten children, scattered children, housewives and the unemployed, workers, commercial service personnel, cadres and clerks, medical staff, farmers, teachers, etc. The top three in terms of distribution were: students, with 637 cases (accounting for 37.30%); kindergarten children, with 258 cases (accounting for 15.11%); and scattered children, with 222 cases (accounting for 13.00%). See Figure 4 for the occupation distribution.

**Fig. 4.**
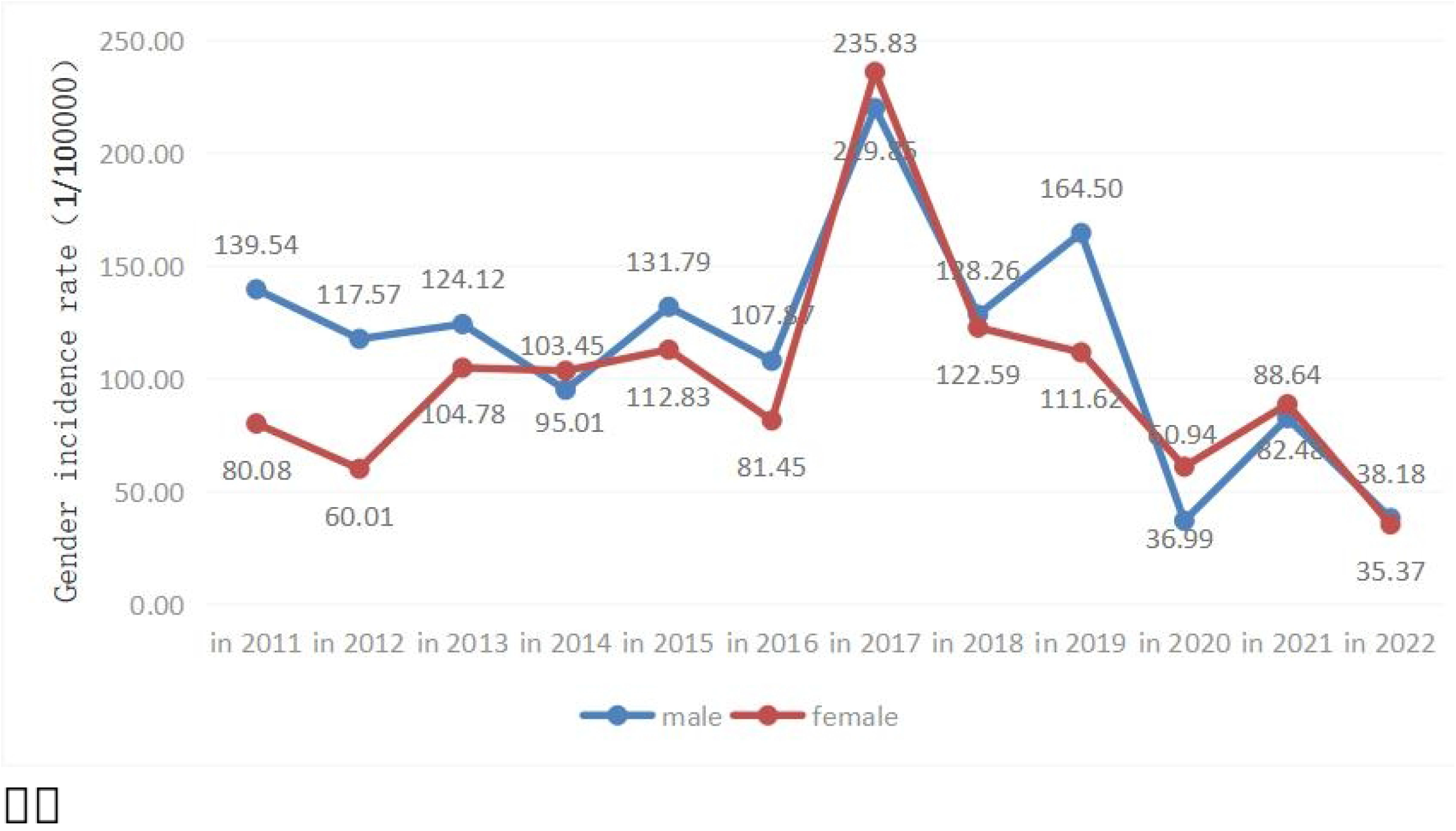

#### Time Distribution

The top three months in terms of cumulative incidence were: November, with 191 cases (accounting for 14.82%); May, with 165 cases (accounting for 12.80%); and July, with 129 cases (accounting for 10.01%). See Figure 5 for the cumulative case time distribution, and Table 1 for the annual case time distribution.

**Fig. 5.**
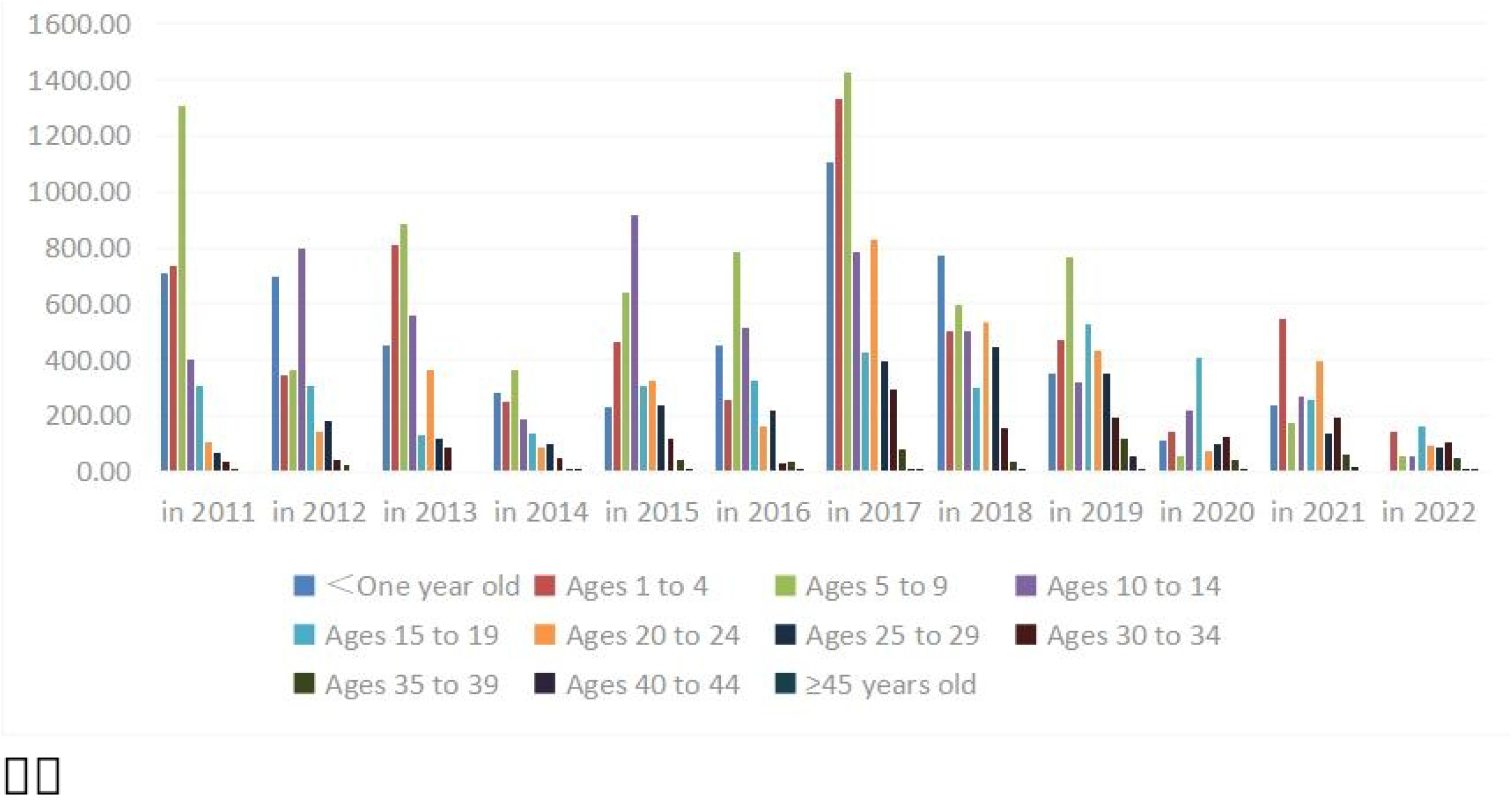

**Table 1.** Temporal distribution of cases per year.

| Year | Jan. | Feb. | Mar. | Apr. | May | Jun. | Jul. | Aug. | Sept. | Oct. | Nov. | Dec. |
| --- | --- | --- | --- | --- | --- | --- | --- | --- | --- | --- | --- | --- |
| 2011 | 8 | 7 | 8 | 4 | 17 | 10 | 11 | 18 | 9 | 10 | 23 | 13 |
| 2012 | 3 | 2 | 5 | 11 | 4 | 5 | 6 | 7 | 5 | 14 | 38 | 13 |
| 2013 | 6 | 7 | 9 | 12 | 25 | 30 | 11 | 14 | 5 | 8 | 15 | 5 |
| 2014 | 2 | 7 | 7 | 21 | 28 | 10 | 14 | 6 | 4 | 0 | 20 | 10 |
| 2015 | 12 | 9 | 11 | 12 | 19 | 17 | 16 | 7 | 13 | 25 | 11 | 9 |
| 2016 | 13 | 7 | 9 | 2 | 14 | 2 | 7 | 10 | 7 | 7 | 22 | 26 |
| 2017 | 14 | 8 | 11 | 27 | 44 | 33 | 36 | 26 | 15 | 28 | 46 | 18 |
| 2018 | 16 | 10 | 11 | 15 | 18 | 12 | 20 | 14 | 10 | 11 | 17 | 16 |
| 2019 | 17 | 11 | 15 | 8 | 9 | 25 | 23 | 16 | 7 | 12 | 21 | 22 |
| 2020 | 2 | 1 | 3 | 4 | 0 | 4 | 4 | 4 | 7 | 8 | 21 | 8 |
| 2021 | 5 | 9 | 6 | 5 | 15 | 7 | 20 | 8 | 7 | 13 | 15 | 8 |
| 2022 | 6 | 4 | 3 | 1 | 0 | 2 | 5 | 4 | 7 | 8 | 5 | 3 |

## 3. Discussion

Varicella is an acute respiratory infectious disease. Although it is not a legally notifiable infectious disease, Shanghai has included varicella case reporting in the China Disease Surveillance Information Reporting Management System since 2005, and a large number of cases are reported every year. The reporting of varicella cases in a suburban area from 2011 to 2022 basically showed a stable wave - like trend, with an obvious increase in 2017, which was slightly different from the incidence situations in the whole country and other provinces and cities ^[7-9]^, but similar to the incidence situations in Pudong New District and Jiading District of Shanghai ^[2, 10]^. It is speculated that the possible reason is the existence of certain regional differences, resulting in different incidence situations.

In terms of gender distribution, both the number of cases and the incidence rate of male patients were higher than those of female patients, which was similar to the situations in other provinces and cities ^[11 - 12]^, but slightly different from the incidence situation in Zhangzhou ^[13]^. The possible reasons are that the total number of males is more than that of females, and females have better hygiene habits than males ^[14]^. Males are more lively in character than females, have more social activities, a wider range of activities, and more exposure to risk factors.

In terms of age distribution, the cases were mainly concentrated in the age group under 14 years old, accounting for 57.20% of all cases, which was basically similar to the age distributions in Jiangsu, Taizhou, Jingzhou, Jiaozhou and other places in China ^[15].^ However, with the passage of time, the incidence rate in the 0 - 14 age group gradually decreased, and the incidence rate in the 15 - 34 age group showed an increasing trend year by year, which was similar to the trends in Jinan and other places in China ^[16]^.

In terms of occupation distribution, the cases were mainly concentrated among students, kindergarten children and scattered children, which was consistent with the age distribution situation and the nature of the disease. The main reason may be that this group of people has a relatively poor immune foundation, has not formed good hygiene habits, is easily infected by the virus and gets sick, and mostly exists in a concentrated manner. In schools and kindergartens, the personnel are dense and the space is limited, which is easy to cause the spread of respiratory infectious diseases.

In terms of time distribution, varicella cases were reported every month in a suburban area from 2011 to 2022. The onset time was basically similar, with obvious peaks, showing a bimodal onset characteristic. There were more cases every year from May to July and in November, which was similar to the research results in Wuhu, Guiyang and other places ^[17 - 18]^, but slightly different from the research results in Zhuhai, Nanjing, Zhangqiu and other places ^[19 - 21]^. The incidence rate showed an obvious peak in 2017. Before 2017, the incidence rate showed a trend of increasing and decreasing every other year, but the overall incidence rate remained stable. From 2020 to 2022, the number of cases decreased significantly. The possible reason is that the COVID - 19 epidemic began in 2020, and Shanghai was under lockdown for 3 months due to the COVID - 19 epidemic in 2022. Residents had a stronger awareness of infectious disease prevention and control and stricter prevention and control measures such as disinfection, resulting in a decrease in the incidence of varicella.

Some studies have shown that if close contacts of varicella cases are vaccinated with varicella vaccine within 5 days, it can effectively prevent the occurrence of the disease or reduce the serious complications caused by varicella ^[22]^. After children are vaccinated with varicella vaccine, with the growth of age, the antibody level in the body gradually decreases. Three years after vaccination, the protective effect of the vaccine decreases significantly, and the incidence rate of varicella breakthrough cases is relatively high ^[23 - 27]^. It is recommended that on the basis of the original planned immunization, a two - dose varicella vaccination program should be implemented, that is, vaccination should be carried out at 12 - 18 months of age, and a second - dose booster immunization should be carried out at 4 - 6 years of age, and the vaccine cost should be borne by parents ^[28 - 30]^. At the end of 2017, Shanghai began to implement the two - needle immunization program for varicella vaccine. Most young children in Shanghai have a two - dose varicella vaccine immunization history. Some literatures show that the second - dose varicella vaccination can make up for the attenuation of the initial immunization protection rate ^[31]^, can significantly reduce the incidence rate of varicella, and reduce the occurrence of varicella epidemics ^[32]^, which is consistent with the decrease in the incidence rate of varicella after 2017.

In view of the above situation and the actual situation of the suburban area, the following suggestions are put forward for the future varicella prevention and control work:

Do a good job in primary prevention. Before the arrival of the annual varicella epidemic peak, corresponding prevention and control measures should be taken in advance, and a joint prevention and linkage system of “hospital - community - school/kindergarten” should be established. After varicella cases are found, measures such as disinfection, isolation and emergency vaccination should be taken in time to reduce the occurrence of clustered epidemics.

Strengthen health education to improve the disease - prevention awareness and knowledge of residents, especially parents of students, so as to prevent diseases in advance.

Strengthen the morning and afternoon inspections and daily disinfection work in schools and kindergartens, improve the monitoring ability and sensitivity, and achieve early detection, early isolation, early treatment and early reporting. In particular, the monitoring and isolation work of the first - onset cases should be done well ^[33]^.

Consolidate the achievements of varicella vaccination, strengthen the promotion of vaccination, and suggest that the government departments adjust the varicella vaccine strategy, include varicella in the national routine immunization program for two - dose vaccination, improve the vaccination rate, and ensure that Children of the appropriate age should receive timely and standardized full-course vaccinations ^[34 - 35]^, so as to reduce the incidence level.

## Data Availability

Data cannot be shared publicly because of the need for information confidentiality and policy requirements.Data are available from the China Disease Prevention and Control Information System for researchers who meet the criteria for access to confidential data.

## Supporting information

**S1 Fig.Varicella case reporting from 2011 to 2022 This figure is used to illustrate the incidence of chickenpox from 2011 to 2022. The horizontal axis represents time and the vertical axis represents the incidence situation. The bar chart indicates the number of cases and the line chart indicates the incidence rate(1/100000)**.

**S2 Fig. Incidence by sex (1/100 000) This figure are used to indicate the incidence rates of different genders (1/100**,**000), with the blue line representing the incidence rate for males and the red line representing that for females**.

**S3 Fig. Incidence by age (1/100 000) This figure is used to indicate the incidence rate (1/ 100**,**000) of different age groups each year. Different colored bars represent different age groups**.

**S4 Fig. Occupational distribution This figure is used to indicate the proportion of different occupations in all cases**.

**S5 Fig. Cumulative case time distribution This figure it is used to show the cumulative case numbers for different months, and different colored lines represent different years**.

**S1 Table Temporal distribution of cases per year This table used to provide detailed information on the number of cases in each month of different years**.

## Notes

### Competing Interest Statement

The authors have declared no competing interest.

### Funding Statement

Yes

### Author Declarations

It has been confirmed by the Ethics Committee of Gaqiao Community Health Service Center, Pudong New Area, Shanghai Municipality that there are no ethical issues, and it is not necessary to seek the consent of individual patients.

